# Higher neighborhood deprivation is associated with accelerated disease progression in behavioral-variant frontotemporal degeneration

**DOI:** 10.1101/2025.05.12.25327099

**Authors:** Rory Boyle, Nadia Dehghani, Sheina Emrani, Anil R. Wadhwani, Melanie Matyi, Katheryn A.Q. Cousins, Emma Rhodes, Brian Nelson, Shana D. Stites, Sharon X. Xie, Laynie Dratch, Vivianna M. Van Deerlin, Allison Snyder, David J. Irwin, Corey T. McMillan, Lauren Massimo

## Abstract

**INTRODUCTION:** Neighborhood deprivation is associated with shorter survival, cognitive impairment and neurodegeneration in aging and Alzheimer’s disease. However, the association of neighborhood deprivation with disease progression in behavioral-variant frontotemporal degeneration (bvFTD) is unknown.

**METHODS:** We examined associations between tertiles of neighborhood deprivation, using the Area Deprivation Index (ADI), and survival in 311 individuals clinically diagnosed with bvFTD from the Penn FTD Center. In a subset (n=161) with complete baseline data across measures of global cognition, executive function, and language, we examined the association of ADI with longitudinal change.

**RESULTS:** Compared to adults living in the least deprived neighborhoods, those living in the most deprived neighborhoods showed shorter survival after symptom onset and faster decline in global cognition, executive and language functions, independent of genetic risk. DISCUSSION: Living in more deprived neighborhoods was associated with an accelerated disease course in bvFTD, highlighting an important socioeconomic disparity in disease prognosis.

**Clinical Trial Registration Information:** N/A

## 1. Introduction

Frontotemporal degeneration (FTD) is a common cause of early-onset dementia with no known cure. It is an understudied neurodegenerative disease that affects the frontal and temporal lobes of the brain and results in progressive deterioration in executive function, language and social comportment. This disease ultimately leads to death but there is considerable variability in survival across pathological subtypes of frontotemporal lobar degeneration (FTLD)^1^. Behavioral variant frontotemporal degeneration (bvFTD) is the most common FTD syndrome^2,3^. There is substantial heterogeneity in the progression of bvFTD across individuals^4^ with a median survival from symptom onset of ∼10 years reported in clinical cohorts^5^, although a lower median survival of ∼6 years is reported in autopsy-confirmed frontotemporal lobar degeneration (FTLD) cases^6^. This heterogeneity presents a major barrier to successful therapeutic interventions given that clinical trial endpoints are challenging to meet without the knowledge of factors that contribute to variable prognosis.

Genetics account for a portion of the heterogeneity in survival and cognitive decline in bvFTD. The presence of a pathogenic (disease-causing) variant in any of the three most common genes associated with autosomal dominant FTD (*MAPT, GRN, C9orf72*) is associated with reduced survival^7^. More rapid cognitive decline is also associated with the presence of a *C9orf72* expansion^8^. Moreover, several common genetic variants including single nucleotide polymorphisms have been associated with reduced survival^9^ and more rapid cognitive decline^10,11^.

Beyond genetics and distinct pathological subgroups of bvFTD, individual-level socioeconomic and lifestyle factors also explain variability in disease progression in bvFTD^6,12,13^ and are therefore thought to contribute to resilience, i.e. better-than-expected cognitive or clinical outcomes given the degree of neurodegenerative disease^14,15^. Better-than-expected outcomes given the degree of genetic risk for neurodegenerative disease also provides evidence for resilience, as seen in Alzheimer’s disease (AD), where slower cognitive decline in apoliprotein-E(*<u>APOE</u>*)-ε4 carriers is associated with higher education and literacy levels^16^. Although resilience is somewhat understudied in FTD compared to AD, there is evidence for protective effects of education, occupation, and leisure activities in FTD^17^. For instance, higher educational and occupational attainment have been associated with better cognitive performance^12^ and longer survival in FTD^6^.

An additional, under-explored source of heterogeneity in bvFTD is neighborhood socioeconomic deprivation, which has been linked to variability in cognitive aging and neurodegenerative disease^18^. Living in socioeconomically deprived neighborhoods can impair access to quality food, education, and healthcare, and is associated with increased exposure to harmful environmental factors^19^, increased stress and worse health outcomes^20,21^, including earlier mortality^22^. The relative deprivation of a neighborhood can be quantified using composite indices, such as the Area Deprivation Index (ADI), which summarizes neighborhood-level information on education, employment, housing, and poverty, collected from the American Community Survey^20,23^. A systematic review of 15 studies reported that higher neighborhood deprivation was associated with worse cognitive function in older adults^24^, which has been further confirmed in several large international cohorts^25–28^. Moreover, neighborhood deprivation is associated with an increased risk of dementia^29,30^, worse late-life cognitive function^31^, shorter survival following dementia diagnosis^32^, faster cognitive decline^33^, and faster cortical thinning in AD-relevant regions, including the entorhinal, precuneus, and middle temporal regions^33^. Recently, we demonstrated that higher neighborhood deprivation, measured with the ADI, is associated with worse late-life cognitive function, independent of the effects of various neurodegenerative proteinopathies^31^. This suggests that neighborhood deprivation contributes to the heterogeneity in neurodegenerative disease outcomes. While growing evidence suggests that neighborhood deprivation is detrimental for cognitive function and dementia-related outcomes, the association of neighborhood deprivation with disease severity and progression in bvFTD has not yet been studied.

In line with previous work^34^, we anchor this study to a bioecological framework^35^ in which ageing processes occur within a complex system of several interconnected levels, including the individual and neighborhood environment. As such, we investigate the hypothesis that greater neighborhood deprivation, measured by the ADI, is associated with an accelerated disease course, characterized by shorter survival and faster cognitive decline, independent of individual-level socioeconomic status, represented by education.

## 2. Methods

### 2.1 Participants

We conducted secondary data analysis of existing data from 311 individuals from the Penn FTD Center (mean age at symptom onset = 59.75 years [SD = 8.89 years], 128 female participants/183 male participants, see Table 1) who met published clinical criteria for probable bvFTD^36^ confirmed by a multidisciplinary consensus committee, and had complete data for the Area Deprivation Index (ADI), date of symptom onset, years of education and valid data for survival analysis were included in this study (see flow chart in e-Fig. 1). Compared to participants with a bvFTD diagnosis who were excluded from this study, included participants were more likely to be older at symptom onset and to have a shorter duration from symptom onset to initial visit, and were less likely to have a confirmed genetic etiology or a co-occurring motor syndrome (see e-Table 1). These participants had their initial visit at the Penn FTD Center between 1994 to 2021 with reported FTD symptom onset ranging from 1985 to 2020. The study period for this analysis ended on 1^st^ May 2024, the date at which clinical data was obtained from the Penn INDD. We define age at symptom onset as an individual’s age in which their symptoms were first noted (by themselves or their caregivers). These 311 individuals formed the group used in the survival analysis (hereafter referred to as the Survival group).

**Table 1.**
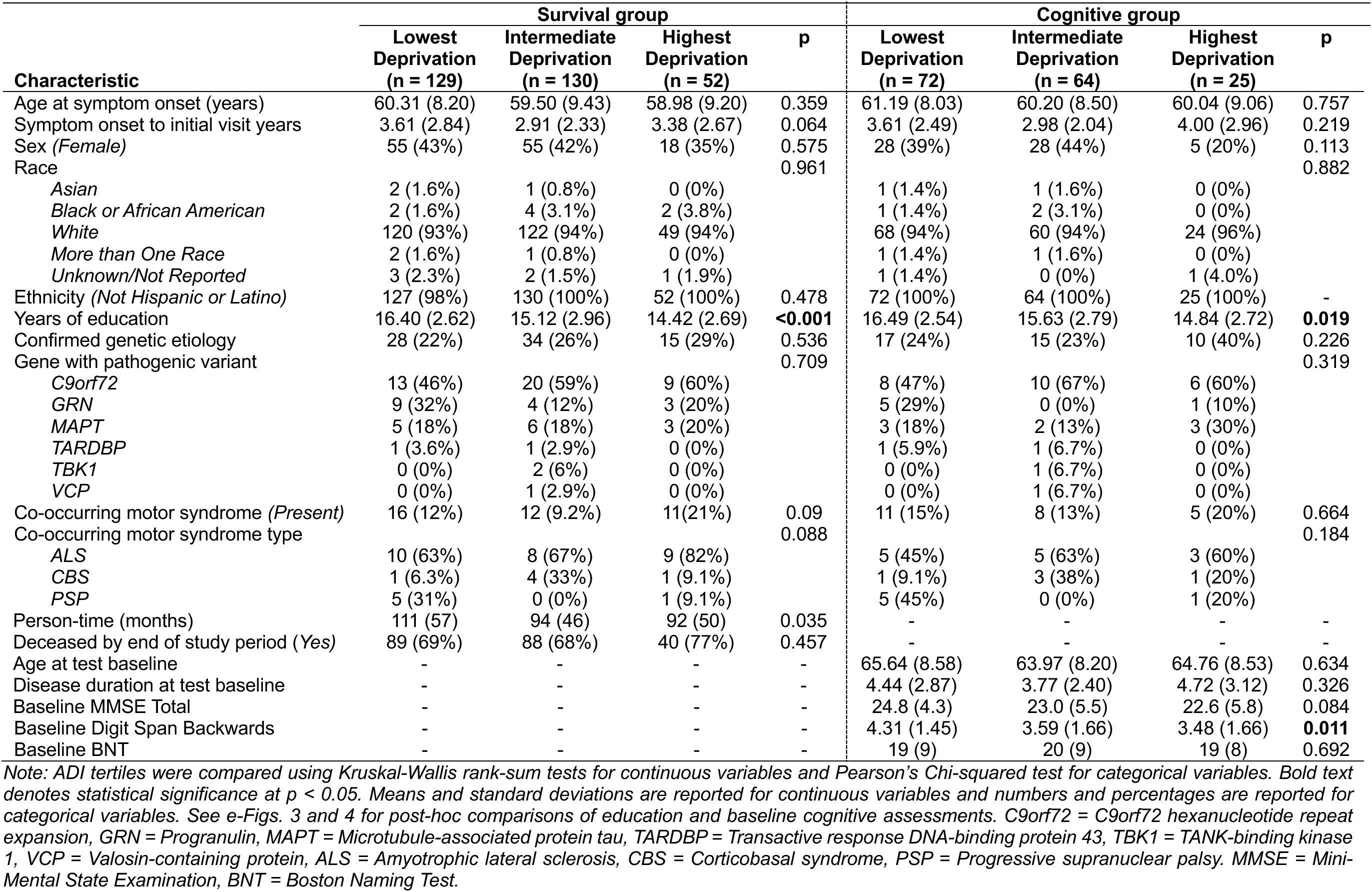
Baseline characteristics of groups used in the survival and cognitive analyses.

| Characteristic | Survival group |  |  |  | Cognitive group |  |  |  |
| --- | --- | --- | --- | --- | --- | --- | --- | --- |
|  | Lowest Deprivation (n = 129) | Intermediate Deprivation (n = 130) | Highest Deprivation (n = 52) | p | Lowest Deprivation (n = 72) | Intermediate Deprivation (n = 64) | Highest Deprivation (n = 25) | p |
| Age at symptom onset (years) | 60.31 (8.20) | 59.50 (9.43) | 58.98 (9.20) | 0.359 | 61.19 (8.03) | 60.20 (8.50) | 60.04 (9.06) | 0.757 |
| Symptom onset to initial visit years | 3.61 (2.84) | 2.91 (2.33) | 3.38 (2.67) | 0.064 | 3.61 (2.49) | 2.98 (2.04) | 4.00 (2.96) | 0.219 |
| Sex (Female) | 55 (43%) | 55 (42%) | 18 (35%) | 0.575 | 28 (39%) | 28 (44%) | 5 (20%) | 0.113 |
| Race |  |  |  | 0.961 |  |  |  | 0.882 |
| Asian | 2 (1.6%) | 1 (0.8%) | 0 (0%) |  | 1 (1.4%) | 1 (1.6%) | 0 (0%) |  |
| Black or African American | 2 (1.6%) | 4 (3.1%) | 2 (3.8%) |  | 1 (1.4%) | 2 (3.1%) | 0 (0%) |  |
| White | 120 (93%) | 122 (94%) | 49 (94%) |  | 68 (94%) | 60 (94%) | 24 (96%) |  |
| More than One Race | 2 (1.6%) | 1 (0.8%) | 0 (0%) |  | 1 (1.4%) | 1 (1.6%) | 0 (0%) |  |
| Unknown/Not Reported | 3 (2.3%) | 2 (1.5%) | 1 (1.9%) |  | 1 (1.4%) | 0 (0%) | 1 (4.0%) |  |
| Ethnicity (Not Hispanic or Latino) | 127 (98%) | 130 (100%) | 52 (100%) | 0.478 | 72 (100%) | 64 (100%) | 25 (100%) | - |
| Years of education | 16.40 (2.62) | 15.12 (2.96) | 14.42 (2.69) | **<0.001** | 16.49 (2.54) | 15.63 (2.79) | 14.84 (2.72) | **0.019** |
| Confirmed genetic etiology | 28 (22%) | 34 (26%) | 15 (29%) | 0.536 | 17 (24%) | 15 (23%) | 10 (40%) | 0.226 |
| Gene with pathogenic variant |  |  |  | 0.709 |  |  |  | 0.319 |
| C9orf72 | 13 (46%) | 20 (59%) | 9 (60%) |  | 8 (47%) | 10 (67%) | 6 (60%) |  |
| GRN | 9 (32%) | 4 (12%) | 3 (20%) |  | 5 (29%) | 0 (0%) | 1 (10%) |  |
| MAPT | 5 (18%) | 6 (18%) | 3 (20%) |  | 3 (18%) | 2 (13%) | 3 (30%) |  |
| TARDBP | 1 (3.6%) | 1 (2.9%) | 0 (0%) |  | 1 (5.9%) | 1 (6.7%) | 0 (0%) |  |
| TBK1 | 0 (0%) | 2 (6%) | 0 (0%) |  | 0 (0%) | 1 (6.7%) | 0 (0%) |  |
| VCP | 0 (0%) | 1 (2.9%) | 0 (0%) |  | 0 (0%) | 1 (6.7%) | 0 (0%) |  |
| Co-occurring motor syndrome (Present) | 16 (12%) | 12 (9.2%) | 11(21%) | 0.09 | 11 (15%) | 8 (13%) | 5 (20%) | 0.664 |
| Co-occurring motor syndrome type |  |  |  | 0.088 |  |  |  | 0.184 |
| ALS | 10 (63%) | 8 (67%) | 9 (82%) |  | 5 (45%) | 5 (63%) | 3 (60%) |  |
| CBS | 1 (6.3%) | 4 (33%) | 1 (9.1%) |  | 1 (9.1%) | 3 (38%) | 1 (20%) |  |
| PSP | 5 (31%) | 0 (0%) | 1 (9.1%) |  | 5 (45%) | 0 (0%) | 1 (20%) |  |
| Person-time (months) | 111 (57) | 94 (46) | 92 (50) | 0.035 | - | - | - | - |
| Deceased by end of study period (Yes) | 89 (69%) | 88 (68%) | 40 (77%) | 0.457 | - | - | - | - |
| Age at test baseline | - | - | - | - | 65.64 (8.58) | 63.97 (8.20) | 64.76 (8.53) | 0.634 |
| Disease duration at test baseline | - | - | - | - | 4.44 (2.87) | 3.77 (2.40) | 4.72 (3.12) | 0.326 |
| Baseline MMSE Total | - | - | - | - | 24.8 (4.3) | 23.0 (5.5) | 22.6 (5.8) | 0.084 |
| Baseline Digit Span Backwards | - | - | - | - | 4.31 (1.45) | 3.59 (1.66) | 3.48 (1.66) | **0.011** |
| Baseline BNT | - | - | - | - | 19 (9) | 20 (9) | 19 (8) | 0.692 |

Note: ADI tertiles were compared using Kruskal-Wallis rank-sum tests for continuous variables and Pearson's Chi-squared test for categorical variables. Bold text denotes statistical significance at $p < 0.05$ . Means and standard deviations are reported for continuous variables and numbers and percentages are reported for categorical variables. See e-Figs. 3 and 4 for post-hoc comparisons of education and baseline cognitive assessments. C9orf72 = C9orf72 hexanucleotide repeat expansion, GRN = Progranulin, MAPT = Microtubule-associated protein tau, TARDBP = Transactive response DNA-binding protein 43, TBK1 = TANK-binding kinase 1, VCP = Valosin-containing protein, ALS = Amyotrophic lateral sclerosis, CBS = Corticobasal syndrome, PSP = Progressive supranuclear palsy. MMSE = Mini-Mental State Examination, BNT = Boston Naming Test.

A subset of the Survival group had complete data across all three selected measures of cognitive function at cognitive test baseline (n = 161, mean age at symptom onset = 60.62 years [SD = 8.34 years], mean age at cognitive test baseline = 64.84 years [SD = 8.40 years], 61 female participants/100 male participants). This subset formed the group used for cognitive analyses (see Table 1, hereafter referred to as the Cognitive group). The cognitive test baseline was anchored to the earliest Mini-Mental State Examination (MMSE) observation collected after year of symptom onset where there was also available data, within 3 months of MMSE, for two other cognitive tests: the Digit Span Backwards and Boston Naming Test (BNT). For the Cognitive group, we define disease duration at test baseline as the time from symptom onset to their baseline cognitive test and the mean disease duration at baseline was 4.22 years [SD = 2.74 years].

**Table 2.** Results of Cox proportional hazards model for survival across ADI tertiles, adjusted for age at onset, sex, education, and genetic status.

| Characteristic | Hazard Ratio | 95% CI | p |
| --- | --- | --- | --- |
| ADI (Intermediate Deprivation (T2)) | 1.36 | 0.99 – 1.88 | 0.057 |
| ADI (Highest Deprivation (T3)) | 1.63 | 1.09 – 2.45 | **0.018** |
| Age at onset | 1.03 | 1.01 – 1.04 | **0.002** |
| Sex (Male) | 0.85 | 0.64 – 1.13 | 0.264 |
| Education | 0.99 | 0.94 – 1.04 | 0.638 |
| Genetic status (Confirmed genetic etiology) | 1.74 | 1.29 – 2.35 | **<0.001** |

*Note: Bold text denotes statistical significance at $p < 0.05$ . The reference groups for the categorical variables ADI, sex, genetic status, and ADI were lowest deprivation tertile, female participants, and no known genetic etiology, respectively. This model provides estimates of the association of each variable with survival, adjusted for other variables in the model.*

All participants or their proxies completed a written informed consent procedure in accordance with the Declaration of Helsinki and approved by the Institutional Review Board of the University of Pennsylvania.

### 2.2 Area deprivation index

We assessed neighborhood deprivation using the ADI, which is a summary index of variables reflecting area-level income, education, employment, and housing quality characteristics^20^. Using 12-digit Federal Information Processing Standard (FIPS) codes from participants’ first clinical visit, we obtained ADI values using the Neighborhood Atlas® website (https://www.neighborhoodatlas.medicine.wisc.edu/). We used the 2020 ADI (version 4) as this variable had the most non-missing data in our cohort. The 2020 ADI was created using 2016-2020 American Community Survey (ACS) 5-Year Data. The ADI provides a national percentile ranking of neighborhoods, at the level of the Census block group, where 1 represents the lowest level of disadvantage within the nation and 100 represents the highest level of disadvantage. Neurodegenerative disease cohorts studies tend to overrepresent lower ADI individuals compared to the broader population^37^ resulting in a skewed ADI distribution. Additionally, the negative effects of neighborhood deprivation are most evident at higher levels of relative deprivation^33^. As such, following previously published methods^31,38^, we categorized the ADI national percentile rank values into tertiles based on the distribution of the ADI across the wider Penn Integrated Neurodegenerative Disease Database^39^ (INDD, n = 20,569) to obtain broadly representative tertiles. The tertiles obtained from INDD were as follows: lowest deprivation (median ADI = 11, range = 1-23); intermediate deprivation (median ADI = 35, range = 24-49); and highest deprivation (median ADI = 62, range = 50-92). Applying these tertiles to the Survival group resulted in 129 individuals in the lowest deprivation (median ADI = 11, range = 1 – 23), 130 in the intermediate deprivation (median ADI = 35, range = 24 – 49), and 52 in the highest deprivation tertile (median ADI = 62, range = 50 – 92). Applying these tertiles to the Cognitive group resulted in 72 individuals in the lowest deprivation (median ADI = 12, range = 2 – 23), 64 in the intermediate deprivation (median ADI = 38, range = 24 – 48), and 25 in the lowest deprivation tertile (median ADI = 57, range = 51 – 81). Although raw ADI values can be used as a study independent method of creating ADI groups (e.g. lowest tertile = ADI 1-33, intermediate tertile = ADI 34-66, highest tertile = ADI 67-100), the skewed distribution of ADI in neurodegenerative disease cohorts can result in very small sample sizes in the highest deprivation tertile. Therefore, we grouped our sample into ADI tertiles based on the distribution across INDD.

### 2.3 Cognitive assessments

While participants in ongoing research in the Penn FTD Center complete an array of cognitive tests, to maximize our sample size and reduce multiple comparisons, we limited our analyses to three tests, collected consistently over a follow-up period spanning 9 years in some individuals, in cognitive domains that are often impaired in bvFTD (global cognition, executive function, and language function).

The MMSE is a screening tool for cognitive impairment^40^ and, although known to be more sensitive to amnestic syndromes, we use it here as a measure of global cognitive performance. MMSE total scores have a possible range of 0 to 30, with higher scores reflecting better global cognitive performance. We had 517 MMSE observations in the Cognitive group, with a minimum interval of 3 months between observations (see Supplement e-Fig. 2 for spaghetti plots of cognitive test scores over time and frequency of observations on each test over time).

We also evaluated two domain-specific cognitive measures. Digit Span Backwards is a working-memory based measure of executive function^41^. Participants are presented with a series of numbers and asked to repeat the series backwards, with increasing span lengths. We used the maximal backwards span, with a possible range of 0 to 8, here as the Digit Span Backwards score. We had available data for 364 Digit Span Backwards observations in the Cognitive group, with a minimum interval of 3 months between observations. The BNT assesses language function, specifically visual naming ability^42^. In this task patients are asked to name stimuli, one-at-time, shown in a picture. We used a 30-item version of the BNT with a possible range of 0 to 30, with 30 representing each stimuli being correctly named. To maximize our number of observations with data on language function over time, and because the BNT is suboptimal for assessing naming ability in non-native English speakers^43^, we also included data collected using the Multilingual Naming test (MINT)^43^. The 32-item MINT is scored with a possible range of 0-32, with 32 representing maximal performance. MINT scores were converted to BNT scores in line with the National Alzheimer’s Coordinating Center (NACC) Crosswalk Study^44^. After conversion, we had available data for 374 BNT observations (145 observations collected using the BNT and 229 collected using the MINT) in the Cognitive group, with a minimum interval of 3 months between observations.

### 2.4 Genetic screening

We determined whether participants had identifiable pathogenic variants in the genes most commonly implicated in hereditary FTD: *C9orf72*, *GRN*, *MAPT*, *TARDBP, TBK1,* and *VCP*^7^. Briefly, DNA was extracted from peripheral blood or frozen brain tissue. *GRN*, *MAPT*, *TARDBP*, *TBK1* and *VCP* mutations were assessed using a custom-targeted next-generation sequencing panel (MiND-Seq) or Exome Sequencing as previously described^45^. *C9orf72* hexanucleotide repeat expansions were assessed using repeat-primed PCR as previously described^46^. In the Survival group, 77 (25%) participants were identified to have a genetic etiology (16 *GRN*, 14 *MAPT*, 2 *TARDBP*, 2 *TBK1*, 1 *VCP*, and 42 *C9orf72*). In the Cognitive group 42 (26%) participants were identified to have a genetic etiology (6 *GRN*, 8 *MAPT*, 2 *TARDBP*, 1 *TBK1*, 1 *VCP*, and 24 *C9orf72*). All other participants were considered to have an unknown genetic status.

### 2.5 Statistical analyses

#### 2.5.1 Survival analysis

Survival time was calculated as the time from symptom onset to death. We included individuals who were still alive at study close, defined as the date (May 1^st^ 2024) at which data were obtained from INDD for analysis, and censored these individuals (n = 94, 30% of Survival group) at their last clinical visit. We additionally adjusted for left truncation in our survival models, which acknowledges the possibility that some individuals with very short survival from symptom onset may not have lived long enough to have entered the study^7^. We accounted for this by considering our data to be left-truncated at the date of initial visit, i.e. the baseline for the survival analysis. We evaluated the difference in survival time from symptom onset across ADI tertiles in a Kaplan-Meier analysis. We conducted the Kaplan-Meier analysis and obtained survival curves with risk tables using the *coxph* and *survfit* functions from the *survival* package in R (version 4.3.2). We then performed Cox regression to examine the association between ADI (reference group = Lowest Deprivation) and survival time (time from symptom onset to death), adjusting for age at symptom onset, sex (reference group = females), years of education, and genetic status (binary variable reflecting the presence of a pathogenic variant, reference group = no known genetic etiology identified). In a sensitivity analysis to account for the shorter survival due to motor symptoms^47^, and in line with previous work^48^, we excluded 39 individuals with a co-occurring diagnosis of either amyotrophic lateral sclerosis (ALS, n = 27), corticobasal syndrome (CBS, n = 6), or progressive supranuclear palsy (PSP, n = 6).

#### 2.5.2 Longitudinal analyses

In separate linear mixed effects models for each cognitive outcome, we examined the association between ADI (reference group = lowest deprivation tertile) and change in cognitive performance over time, adjusting for age at baseline test, sex (reference group = female participants), years of education, baseline test score, disease duration at baseline, and genetic status (reference group = no known genetic etiology identified). Individual was included as a random intercept term in each model. Our effect of interest in each linear mixed effects model was the time from baseline x Highest Deprivation tertile interaction term. We conducted the linear mixed effect models using the *lme* function from the *nlme* package in R.

## 3. Results

### 3.1 Participant characteristics

Participant characteristics of the Survival and Cognitive groups are shown in Table 1. 25% of the Survival group and 26% of the Cognitive group had confirmed genetic etiology (the presence of an FTD-causing pathogenic variant, see Section 2.4 Genetic screening). 13% of the Survival group and 15% of the Cognitive group had co-diagnoses of FTLD-associated motor syndromes. The demographics of the Cognitive group did not differ from the Survival group (all p>.05, see Supplement e-Table 2). In the Survival group, the intermediate and highest ADI tertiles had significantly fewer years of education than the lowest ADI tertile (see e-Fig. 3). In the Cognitive group, the highest ADI tertile had significantly fewer years of education than the lowest ADI tertile (see e-Fig. 3). At baseline, the intermediate and highest ADI tertiles had significantly worse Digit Span Backwards performance than the lowest ADI tertiles, but baseline performance on MMSE and BNT was not significantly different across ADI tertiles (see e-Fig. 4 for boxplots of baseline cognitive assessments). There were no statistically significant differences across ADI tertiles in other participant characteristics (see Table 1). The lowest ADI tertile had a significantly greater number of observations than the intermediate and highest ADI tertiles on average for the MMSE but there were no significant group differences in number of observations across ADI tertiles for Digit Span Backwards and BNT (see e-Fig. 2).

### 3.2 Higher neighborhood deprivation is associated with shorter survival in bvFTD

A Kaplan-Meier analysis indicated that higher ADI was associated with shorter survival (see Fig. 1). This association of ADI with survival was confirmed in a Cox proportional hazards model (n = 311, events = 217, censored = 94) adjusting for age at symptom onset, sex, years of education, and genetic status, where, compared to the lowest ADI tertile as a reference group, the highest ADI tertile showed shorter survival (hazard ratio = 1.63, 95% CI = 1.09 – 2.45, p = 0.018, see Table 2). The intermediate ADI tertile also showed shorter survival compared to the lowest ADI tertile, but this association was not statistically significant (hazard ratio = 1.36, 95% CI = 0.99 – 1.88, p = 0.057). Older age at symptom onset and having a confirmed genetic etiology were also associated with shorter survival. Sex and years of education were not associated with shorter survival when adjusting for age at symptom onset, ADI, and genetic status. The highest ADI tertile did not significantly differ, compared to the lowest ADI tertile, in time between symptom onset and initial visit (see Table 1), suggesting that these findings are not attributable to a diagnostic delay.

**Figure 1.**
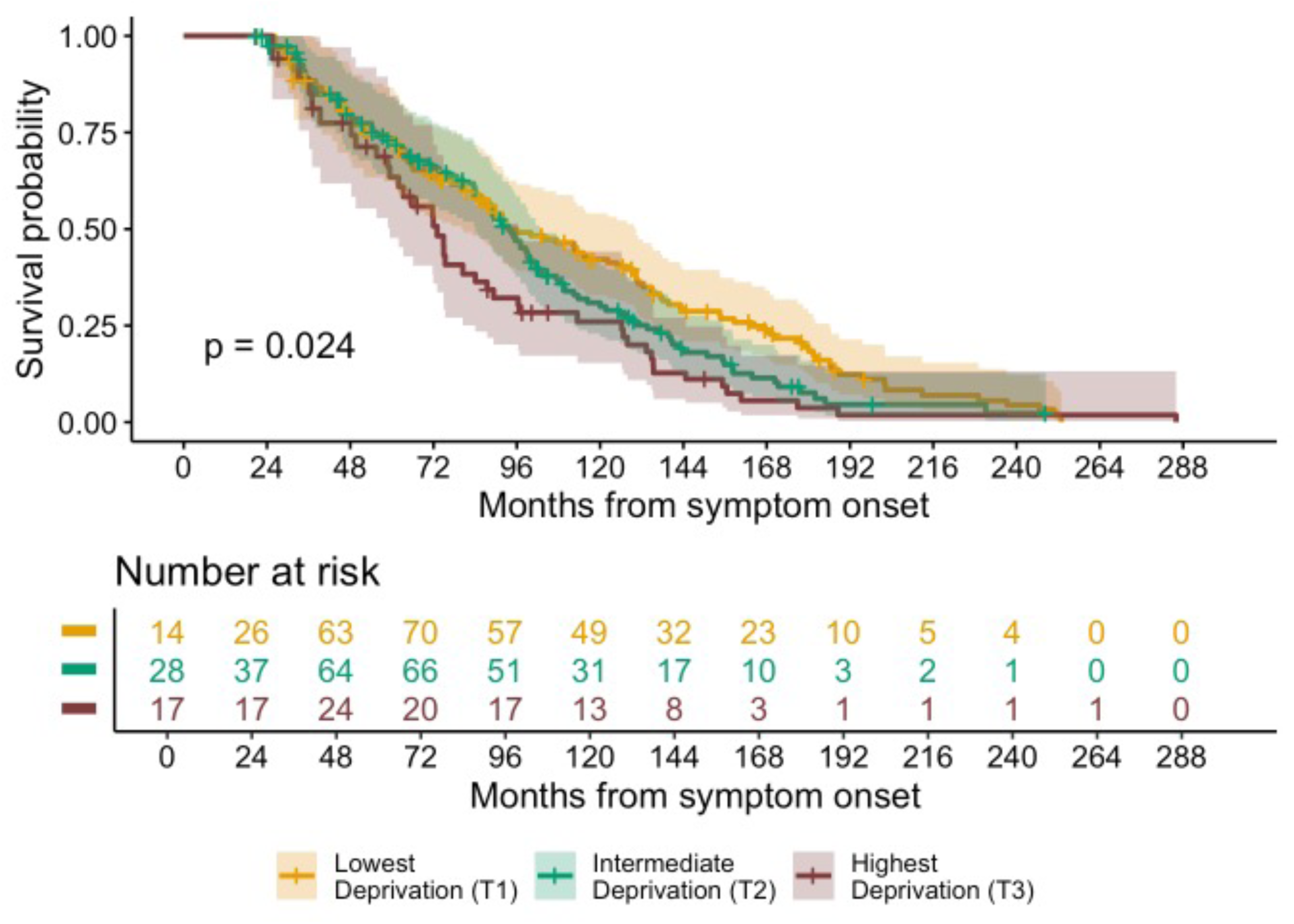
Kaplan-Meier curves with log-rank p-value showing survival from symptom onset across ADI tertiles. Note: Because of delayed entry due to left-truncation in the Cox model, the risk set size increases after time 0 (i.e. 0 months from symptom onset).

Sex-specific median survival was estimated for those with and without confirmed genetic etiology based on the mean age at symptom onset (59.75 years old) and mean years of education (15.53 years) across the whole sample and decreased in a dose-response type manner with shorter median survival in each increasing tertile of ADI (see Table 3). The Cox proportional hazards model satisfied the assumption of proportional hazards (global χ^2^ = 8.99, p = 0.17).

**Table 3.**
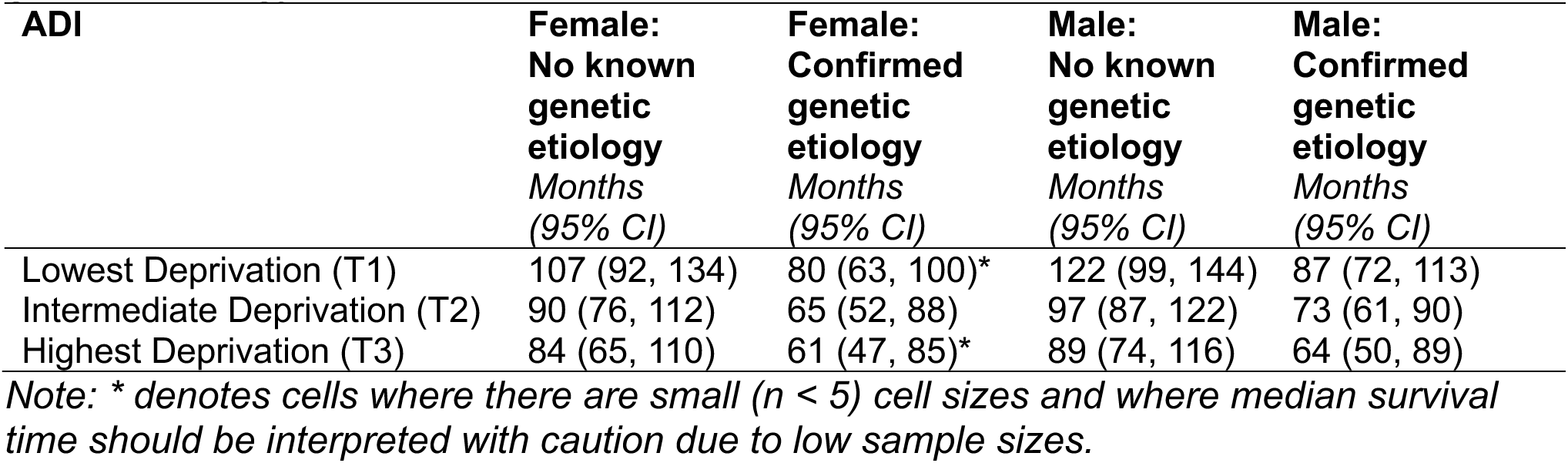
Sex-specific median survival time for those with and without known genetic etiology across ADI tertiles.

In sensitivity analyses, the association of higher ADI with reduced survival remained statistically significant (HR = 1.64, 95% CI = 1.03 – 2.60, p = 0.036) even after removing individuals with co-occurring motor syndromes (n=39, see Table 1) and adjusting for age at onset, sex, education, and genetic status in this subsample (see Supplement e-Fig. 5 and e-Table 3).

### 3.3 Higher neighborhood deprivation is associated with faster cognitive decline in bvFTD

Separate linear mixed effects models, adjusting for sex, education, age at baseline, baseline test performance, disease duration at baseline, and genetic status, revealed that, compared to the lowest ADI tertile, the highest ADI tertile showed faster decline in performance over time (see Figure 2 and Table 4) on the MMSE (p = 0.011), Digit Span Backwards (p = 0.002) and BNT (p = 0.029). Compared to the lowest ADI tertile, there was no statistically significant difference in decline in performance over time in the intermediate ADI tertile on the MMSE (p = 0.148), Digit Span Backwards (p = 0.127) or the BNT (p = 0.563).

**Figure 2.**
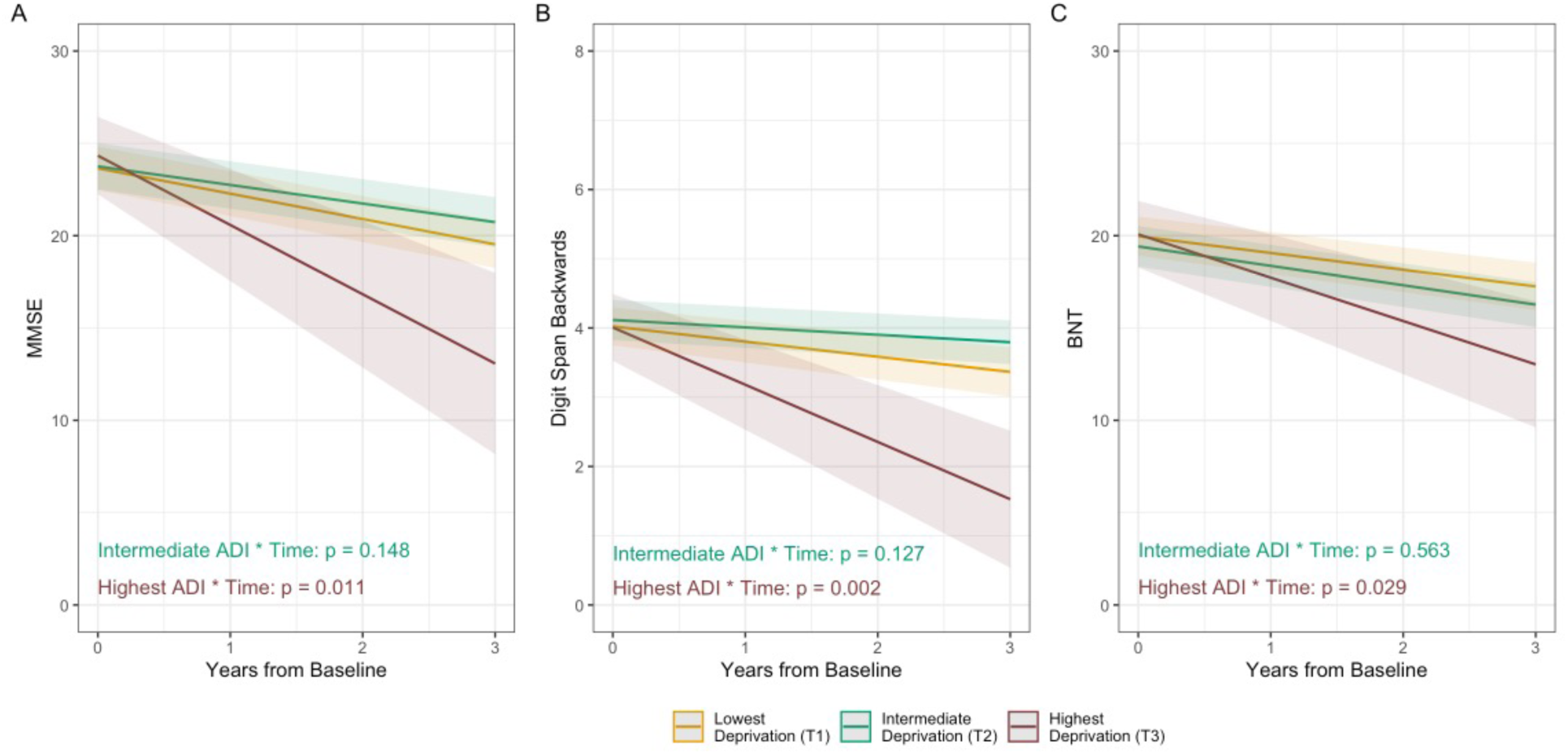
Associations of ADI with change over time in cognitive performance. A) MMSE, B) Digit Span Backwards, C) BNT. *Note: Marginal effects of the ADI tertile x time interaction are plotted here when continuous variables are equal to their mean values, and factor variables equal their reference values (sex = female, genetic status = no known genetic etiology). The effects are estimated from models fit using the full follow-up timeframe but are plotted for the first 3 years of follow-up to improve interpretability as this timeframe had the densest data coverage across deprivation tertiles*.

**Table 4.** Results of linear mixed effects models examining the association of ADI with cognitive change over time.

| Predictors | *MMSE*<br>*N = 161, Observations = 517* |  |  | *Digit Span Backwards*<br>*N = 161, Observations = 364* |  |  | *BNT*<br>*N = 161, Observations = 374* |  |  |
| --- | --- | --- | --- | --- | --- | --- | --- | --- | --- |
|  | *B* | *CI* | *p* | *B* | *CI* | *p* | *B* | *CI* | *p* |
| Intercept | -2.06 | -8.19 – 4.07 | 0.515 | -0.85 | -2.10 – 0.40 | 0.190 | -0.40 | -5.19 – 4.38 | 0.871 |
| Time (from baseline) | -1.37 | -1.72 – -1.02 | **<0.001** | -0.22 | -0.33 – -0.11 | **<0.001** | -0.91 | -1.28 – -0.54 | **<0.001** |
| ADI (Intermediate Deprivation) | 0.11 | -1.24 – 1.46 | 0.875 | 0.09 | -0.22 – 0.41 | 0.568 | -0.56 | -1.72 – 0.61 | 0.356 |
| ADI (Highest Deprivation) | 0.69 | -1.35 – 2.73 | 0.511 | -0.01 | -0.49 – 0.46 | 0.951 | 0.10 | -1.61 – 1.81 | 0.908 |
| Sex (Male) | 0.83 | -0.39 – 2.05 | 0.185 | -0.01 | -0.29 – 0.27 | 0.941 | 1.02 | -0.08 – 2.12 | 0.073 |
| Education | -0.10 | -0.33 – 0.12 | 0.362 | 0.01 | -0.04 – 0.06 | 0.709 | -0.07 | -0.26 – 0.13 | 0.509 |
| Age at test baseline | 0.00 | -0.07 – 0.07 | 0.977 | 0.02 | 0.00 – 0.03 | **0.041** | -0.01 | -0.07 – 0.05 | 0.849 |
| Baseline score | 1.06 | 0.93 – 1.18 | **<0.001** | 0.90 | 0.81 – 0.99 | **<0.001** | 1.01 | 0.96 – 1.07 | **<0.001** |
| Disease duration at test baseline | 0.24 | 0.03 – 0.45 | **0.024** | -0.03 | -0.07 – 0.02 | 0.238 | 0.15 | -0.03 – 0.33 | 0.102 |
| Genetic status (Confirmed genetic etiology) | 0.09 | -1.22 – 1.39 | 0.894 | 0.08 | -0.22 – 0.38 | 0.605 | -0.13 | -1.26 – 1.00 | 0.822 |
| ADI (Intermediate Deprivation) x Time | 0.36 | -0.12 – 0.85 | 0.148 | 0.11 | -0.03 – 0.25 | 0.127 | -0.14 | -0.62 – 0.34 | 0.563 |
| ADI (Highest Deprivation) x Time | -2.38 | -4.20 – -0.56 | **0.011** | -0.61 | -0.99 – -0.23 | **0.002** | -1.44 | -2.71 – -0.17 | **0.029** |

*Note: All models included a random intercept term. The reference groups for the categorical variables ADI, sex, genetic status were lowest deprivation tertile, female participants, and no known genetic etiology, respectively. Bold text denotes statistical significance at $p < 0.05$ .*

## 4. Discussion

In a clinical cohort, we observed that individuals with bvFTD living in neighborhoods with higher levels of deprivation (i.e. above the 50^th^ percentile nationally) had an accelerated disease course. Specifically, higher deprivation was associated with shorter survival and faster decline in global cognition, executive function, and language function. These findings indicate that the neighborhood environment may contribute to the substantial variability seen in bvFTD progression.

Our finding that higher neighborhood deprivation was associated with reduced survival from symptom onset in bvFTD is in line with associations of higher deprivation with reduced survival in other dementias, including Alzheimer’s disease and Parkinson’s disease^32^, and broadly in other diseases such as cancer^49^ or stroke^50^, and with earlier all-cause mortality^51^. We observed a substantial association, as individuals living in the highest tertile of neighborhood deprivation had, on average, between 19 (in females with confirmed genetic etiology) to 33 months (in males without known genetic etiology) shorter median survival compared to individuals in the lowest tertile. Moreover, this association was independent of the effects of age at onset, sex, years of education, genetic status as well as the presence of co-occurring motor syndromes^5,47,52^. This novel and robust association suggests that neighborhood deprivation may contribute to an accelerated disease course in bvFTD.

We found that individuals with bvFTD living in the highest tertile of neighborhood deprivation showed more rapid decline in global cognitive performance, executive function and language function than individuals in the least deprived tertile. Specifically, individuals in the highest deprivation tertile, compared to those in the lowest tertile, displayed a 2.38 point faster annual decline in MMSE scores (equating to a 7.9% greater reduction in total score per year), a 0.61 point faster annual decline on the Digit Span Backwards test (7.6% greater reduction in digit span per year), and 1.44 point faster annual decline on the Boston Naming Test (4.8% greater reduction in overall items named per year). These effects are notable, as the observed effect size for the MMSE is large enough to be considered a clinically meaningful change in AD clinical trials ^53^, based on analysis from the National Alzheimer’s Coordinating Center Uniform Dataset which identified a 1-3 point decrease in MMSE as a clinically meaningful decline. However, given the small number of individuals (n = 25) in the highest deprivation tertile in these analyses and relatively wide confidence intervals, we should be cautious in drawing firm conclusions about the magnitude of these effects.

Our results extend previous findings of associations of neighborhood deprivation with global cognitive decline in a large epidemiologic cohort^54^, with executive function and memory decline in a cohort with dementia^55^, and with executive function decline in a cohort of adults who were cognitively unimpaired at baseline^33^. In the latter cohort, the association between deprivation and executive function decline was partially mediated by cortical thinning in AD-signature regions (i.e. the inferior parietal, inferior temporal, middle temporal, entorhinal, fusiform, and precuneus)^33^. Future work will allow us to understand if the similar association observed in bvFTD is mediated by frontotemporal lobe atrophy.

It is well-established that socioeconomic deprivation is a fundamental cause of negative health-related outcomes^21^ (e.g. multi-morbidity^56^ and earlier all-cause mortality^51^) and reflects access to resources (e.g. money, knowledge, power, social connections) that allow individuals to avoid disease risk factors, engage with protective factors, and minimize the consequences of disease^21^. The fundamental cause of deprivation may underlie and/or exacerbate the other mechanisms, which are not necessarily independent. Beyond this broad negative impact of deprivation on general health outcomes, the contribution of neighborhood deprivation to an accelerated disease course in bvFTD may be explained via several mechanisms.

First, individuals in more deprived neighborhoods have a higher risk of overexposure to heat, pollutants, and lack of vegetation or greenspace^19^. These exposures are associated with increased risk of dementia^57,58^ and dementia-related mortality^59^, and accelerated epigenetic aging^60^. As such, greater neighborhood deprivation may give rise to an accelerated disease course in bvFTD due to biological pathways associated with harmful exposures. The influence of specific exposures on the disease course in bvFTD warrants further investigation.

Second, neighborhood deprivation may also influence outcomes in bvFTD via epigenetic and psychosocial pathways. Data from the Baltimore Memory Study^61^ demonstrated a novel gene-by-environment interaction in AD whereby perceived neighborhood stressors were associated with worse executive function performance but only in *APOE* e4 carriers. Disparities in “epigenetic age acceleration” in the Health and Retirement Study are explained in part by neighborhood deprivation and physical environment exposures^60^ and epigenetic age acceleration is associated with more rapid decline in general cognition and functional outcomes^62^. Higher neighborhood deprivation has been related to differential DNA methylation at a CpG site, cg26514961, located within the *PLXNC1* gene that may be involved in the immune response^63^. This suggests that increased psychosocial stress from living in a more deprived neighborhood could alter immune responses.

Third, individuals living in deprived neighborhoods may have had fewer opportunities to engage in cognitively stimulating activities that strengthen cognitive resilience^64^. Individuals in more deprived neighborhoods may also have been more likely to attend under-resourced schools, which may reduce cognitive resilience given that lower educational quality is associated with increased dementia risk, independently of educational attainment, which we covaried for in our analyses^65^. Consequently, individuals living in deprived areas may be less able to compensate in the face of neurodegenerative disease and pathological processes. In the present study, we observed associations of higher deprivation with faster cognitive decline, independent of the effect of genetic contribution to bvFTD. This suggests that neighborhood deprivation is associated with reduced cognitive resilience to neurodegenerative disease, and is line with post-mortem data showing that greater neighborhood deprivation is associated with worse late-life cognitive function, independent of the effect of various dementia-related neuropathologies^31^.

Our findings suggest that population-level structural interventions are necessary^66^ to reduce socioeconomic inequality and its consequences for neurodegenerative disease outcomes. Such interventions could also strive to ensure greater access to resources that provide opportunities for cognitive stimulation and enrichment (e.g. libraries, community centers, Men’s Sheds^67^, Women’s Sheds^68^) and more equitable provision of high-quality education in more deprived neighborhoods^18^. Healthcare systems should also allocate additional resources to individuals with bvFTD who live in areas of high deprivation as these individuals are at increased risk for worse outcomes. This could include devising strategies to improve affordability and accessibility of care for symptom management, by providing access to nurses and social workers within these high deprivation areas or by providing transport to and from healthcare clinics.

There are some limitations to our findings to consider. We measured survival from symptom onset which is based on inherently subjective information. Accurate recall of symptom onset in bvFTD is particularly challenging as it can involve behaviors or neuropsychiatric symptoms that are not typical of other neurodegenerative disorders. Nonetheless, it is common in FTD research to measure survival from symptom onset^6,7^ and doing so further allowed us to adjust for left-truncation in the survival analysis. The ADI may be largely weighted towards house values and can therefore underestimate deprivation in some urban areas^69^. This is evident in a discordance between the ADI and other deprivation indices in several US cities^70^. However, higher ADI values have been linked to worse cognitive impairment^31^ and other negative health outcomes in the Philadelphia area^71,72^. Future work could attempt to replicate findings with other deprivation indices, e.g. the neighborhood disadvantage index from the National Neighborhood Data Archive (NaNDA)^34^. NaNDA also provides publicly available data of various neighborhood factors that we can study in future work to identify specific factors driving the association with worse outcomes in bvFTD. In our analyses of cognitive change, the high ADI tertile contained only 25 individuals which limited our ability to carry out exploratory subgroup analyses. Developing an understanding of the specific mechanisms driving the associations of deprivation with worse outcomes in bvFTD will require larger sample sizes and a greater representation of individuals from high ADI areas. FTD clinics and research studies could consider using the ADI to identify areas where recruitment efforts and outreach activities could be focused. Given the small sample size of the high ADI tertile, our findings should be replicated in other cohorts to confirm that these associations generalize beyond this cohort. Importantly, we measured ADI at a single point in the individual’s life course which neglects their residential history. Future work using emerging tools that can reconstruct residential histories^73^ may allow us to identify critical periods for the influence of neighborhood deprivation on outcomes in bvFTD. This will also allow us to account for residential transitions from living at home in the community to living in care facilities. Future work using tau and TDP-43 specific biomarkers will enable testing of these associations within pathological subgroups. In the present study, however, we accounted for biological factors including known pathogenic variants as well as co-occurring motor syndromes. Moreover, incorporating epigenetics and plasma and neuroimaging biomarkers will allow us to identify potential biological mechanisms through which neighborhood deprivation may affect progression. Given the prominent behavioral impairments in bvFTD, future work should also evaluate the influence of deprivation on longitudinal behavioral change.

In conclusion, in a longitudinal clinical cohort of individuals with bvFTD, in analyses that adjusted for several potential confounds, we observed that individuals living in the most deprived neighborhoods, compared to those living in the least deprived neighborhoods, displayed an accelerated disease course, characterized by reduced survival and faster cognitive decline.

## Data Availability

Data will be made available upon reasonable request.

https://www.pennbindlab.com/data-sharing

## Acknowledgements

We thank our participants as well as their caregivers and families for the time and effort that they have generously contributed to this research. We would also like to gratefully acknowledge the valuable work and dedication of our colleagues in the Penn FTD Center in collecting this data and for their thoughtful feedback on this work.

