## Supplement for "Higher neighborhood deprivation is associated with accelerated disease progression in behavioral-variant frontotemporal degeneration"

| **Characteristic** | **Excluded** | **Included** | **p** |
|---|---|---|---|
| ADI Tertile* | | | 0.557 |
| *Lowest Deprivation* | 64 (46%) | 129 (41%) | |
| *Intermediate Deprivation* | 51 (36%) | 130 (42%) | |
| *Highest Deprivation* | 25 (18%) | 52 (17%) | |
| ADI National Rank* | 31 (21) | 30 (20) | 0.759 |
| Age at symptom onset (years)* | 57.69 (8.12) | 59.75 (8.89) | **0.048** |
| Symptom onset to initial visit years* | 3.68 (2.39) | 3.28 (2.62) | **0.037** |
| Sex *(Female)* | 79 (47%) | 128 (41%) | 0.196 |
| Race | | | 0.377 |
| *Asian* | 5 (3.0%) | 3 (1.0%) | |
| *Black or African American* | 6 (3.6%) | 8 (2.6%) | |
| *White* | 151 (90%) | 291 (94%) | |
| *More than One Race* | 3 (1.8%) | 3 (1.0%) | |
| *Unknown/Not Reported* | 2 (1.2%) | 6 (1.9%) | |
| Ethnicity *(Not Hispanic or Latino)* | 163 (98%) | 309 (99%) | 0.157 |
| Years of education* | 15.72 (3.02) | 15.53 (2.87) | 0.290 |
| Confirmed genetic etiology | 22 (13%) | 77 (25%) | **0.002** |
| Gene with pathogenic variant | | | 0.514 |
| *C9orf72* | 12 (55%) | 42 (55%) | |
| *GRN* | 5 (23%) | 16 (21%) | |
| *MAPT* | 3 (14%) | 14 (18%) | |
| *TARDBP* | 0 (0%) | 2 (2.6%) | |
| *TBK1* | 0 (0%) | 2 (2.6%) | |
| *VCP* | 1 (4.5%) | 1 (1.3%) | |
| *Huntingtin* | 1 (4.5%) | 0 (0%) | |
| Co-occurring motor syndrome *(Present)* | 9 (5.3%) | 39 (13%) | **0.011** |
| Co-occurring motor syndrome type | | | 0.857 |
| *ALS* | 6 (67%) | 27 (69%) | |
| *CBS* | 1 (11%) | 6 (15%) | |
| *PSP* | 2 (22%) | 6 (15%) | |

**e-Table 1. Comparison of demographics between individuals with a bvFTD diagnosis who were included in the survival analysis and individuals who were excluded.** Exclusion criteria were missing data for the ADI, date of symptom onset, age at symptom onset, or education and/or invalid data for survival analysis. *Denotes variable with missing data. *Bold text denotes statistical significance at p < 0.05. Means and standard deviations are reported for continuous variables and numbers and percentages are reported for categorical variables. C9orf72 = C9orf72 hexanucleotide repeat expansion, GRN = Progranulin, MAPT = Microtubule-associated protein tau, TARDBP = Transactive response DNA-binding protein 43, TBK1 = TANK-binding kinase 1, VCP = Valosin-containing protein, ALS = Amyotrophic lateral sclerosis, CBS = Corticobasal syndrome, PSP = Progressive supranuclear palsy. MMSE = Mini-Mental State Examination, BNT = Boston Naming Test.*

| Characteristic | Test statistic | p |
|---|---|---|
| Age at onset | t = 1.33 | 0.187 |
| Years of education | t = 1.65 | 0.1 |
| Sex | $\chi^2$ = 0.71 | 0.399 |
| ADI Tertiles | $\chi^2$ = 0.15 | 0.702 |
| Known genetic etiology | $\chi^2$ = 0.82 | 0.696 |

**e-Table 2. Comparison of demographics between Survival and Cognitive groups.** One-sample t-tests compared the mean age at onset and years of education in the Cognitive group to the overall mean (i.e. mean of the Survival group). Chi-squared tests compared the frequencies of categorical variables in the Cognitive group to the overall Survival group.

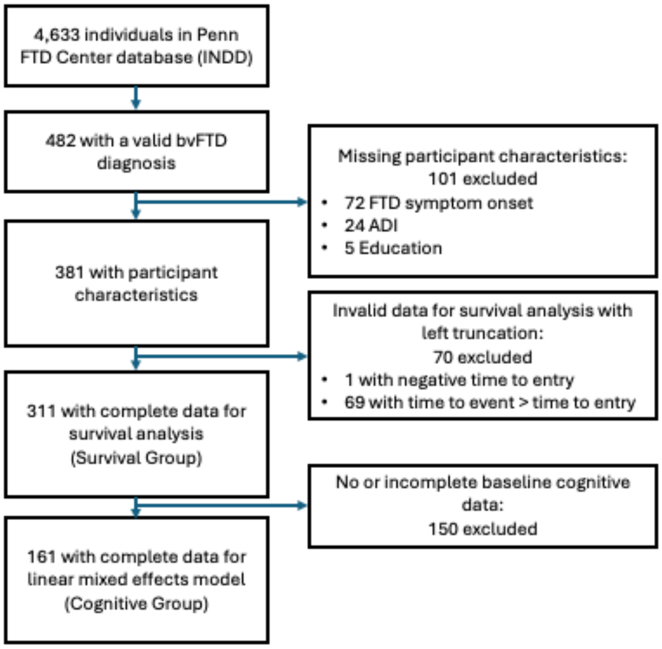


**eFigure 1. Participant flow chart for Survival and Cognitive groups.**

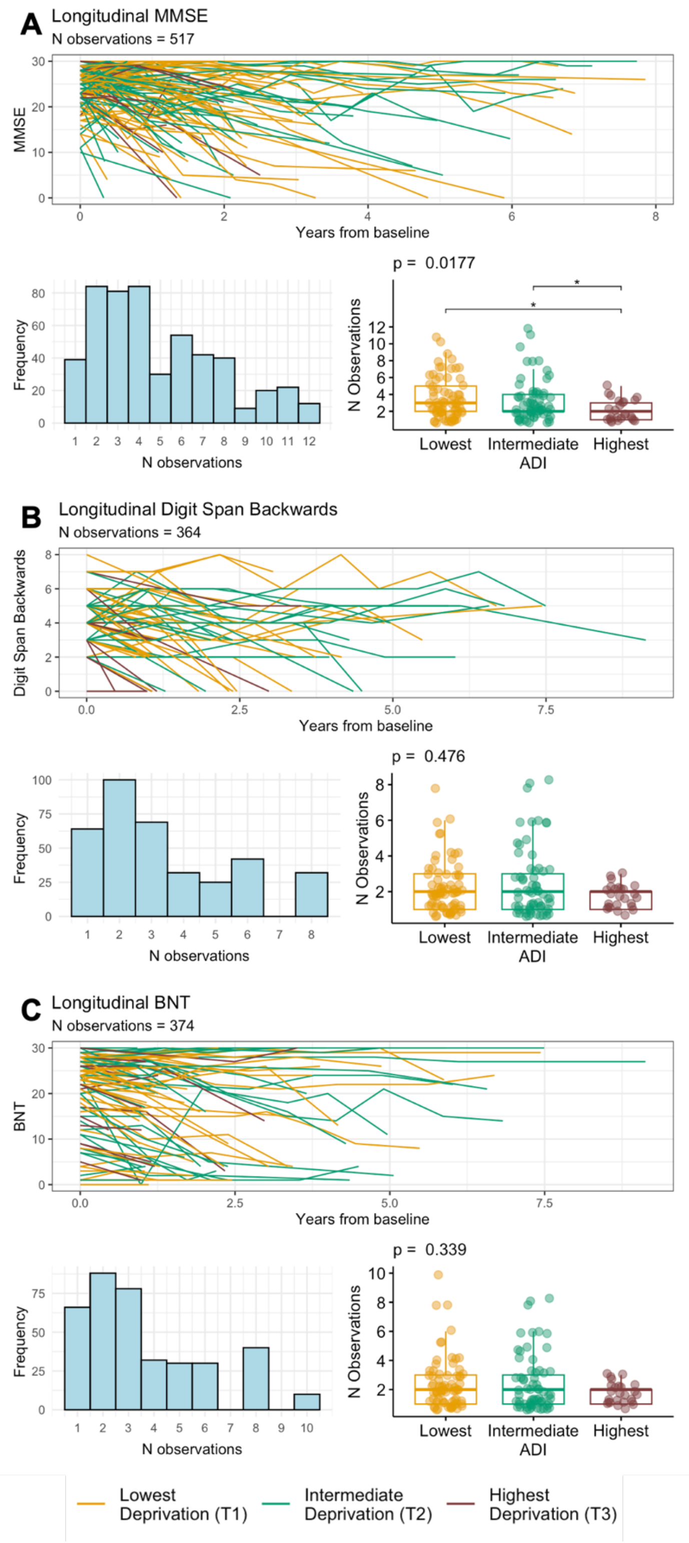


**eFigure 2. Spaghetti plots of cognitive assessments over time and frequency of observations.** Number of observations were compared across ADI tertiles using non-parametric Kruskal-Wallis tests with Dunn's post hoc tests for pairwise comparisons, corrected for multiple comparisons using the Hochberg method.

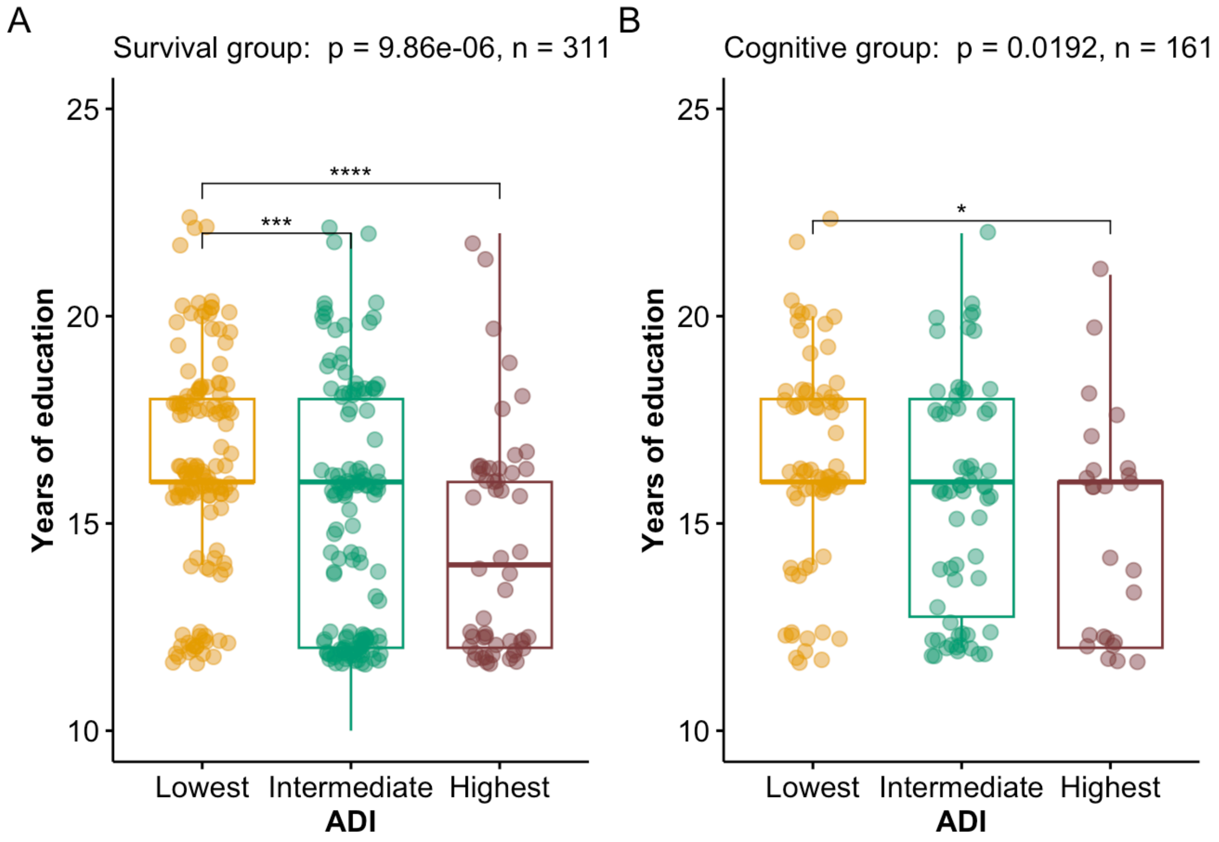


**e-Figure 3. Years of education across ADI tertiles in A) Survival group and B) Cognitive group.** Years of education were compared across ADI tertiles using non-parametric Kruskal-Wallis tests with Dunn's post hoc tests for pairwise comparisons, corrected for multiple comparisons using the Hochberg method.

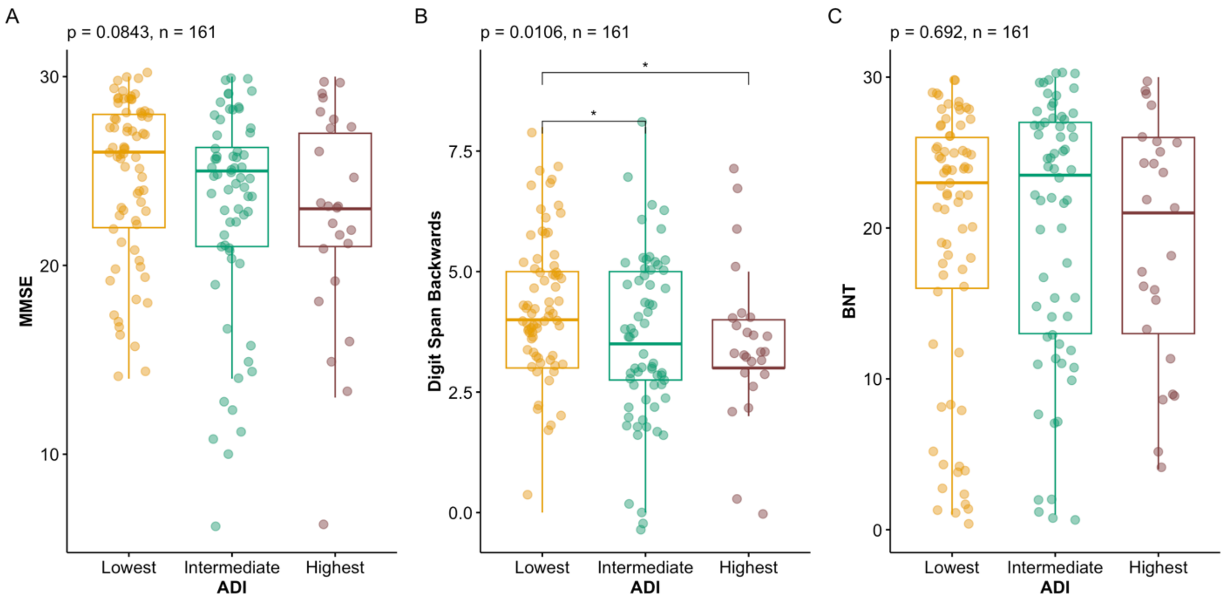


**e-Figure 4. Baseline cognitive assessments across ADI tertiles.** Cognitive assessments were compared across ADI tertiles using non-parametric Kruskal-Wallis tests with Dunn's post hoc tests for pairwise comparisons, corrected for multiple comparisons using the Hochberg method.

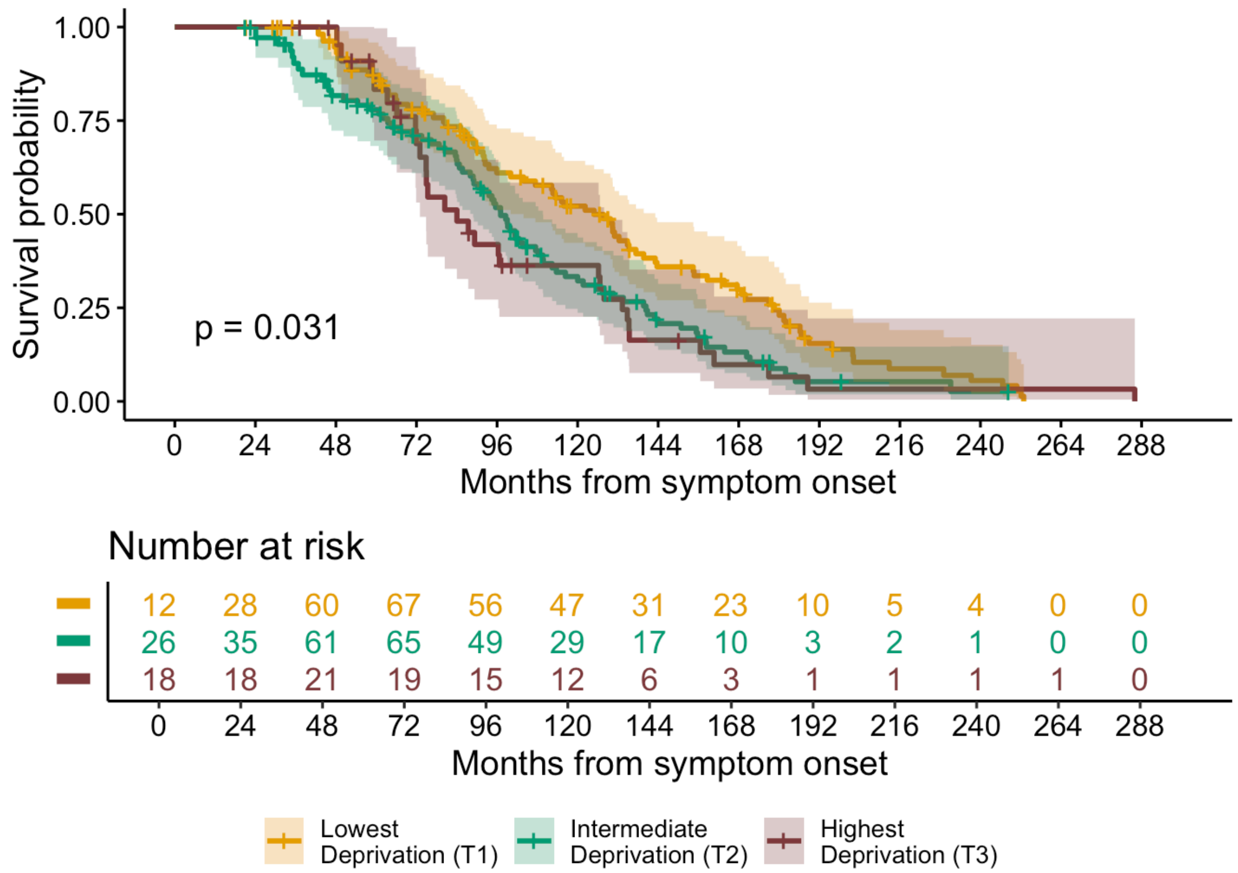


**e-Figure 5. Kaplan-Meier curves with log-rank p-value showing survival from symptom onset across ADI tertiles excluding individuals with co-occurring motor syndromes.** *Note: Because of delayed entry due to left-truncation in the Cox model, the risk set size increases after time 0 (i.e. 0 months from symptom onset).*

**e-Table 3. Results of Cox proportional hazards model for survival across ADI tertiles, excluding individuals with co-occurring motor syndromes, adjusted for age at onset, sex, education, and genetic status.**

| **Characteristic** | **Hazard Ratio** | **95% CI** | **p** |
|---|---|---|---|
| ADI (Intermediate Deprivation (T2)) | 1.55 | 1.10, 2.20 | **0.013** |
| ADI (Highest Deprivation (T3)) | 1.64 | 1.03, 2.60 | **0.036** |
| Age at onset | 1.03 | 1.02, 1.05 | **<0.001** |
| Sex (Male) | 0.82 | 0.60, 1.12 | 0.208 |
| Education | 1.00 | 0.94, 1.05 | 0.878 |
| Genetic status (Confirmed genetic etiology) | 1.68 | 1.21, 2.33 | **0.002** |

*Note: Bold text denotes statistical significance at $p < 0.05$. The reference groups for the categorical variables ADI, sex, genetic status, and ADI were lowest deprivation tertile, female participants, and no known genetic etiology, respectively. The Cox proportional hazards model satisfied the assumption of proportional hazards (global $\chi^2 = 4.93$, $p = 0.55$).*
